# The effects of nicotine on neurocognitive outcomes: evidence from a polygenic risk score analysis

**DOI:** 10.64898/2025.11.27.25341175

**Authors:** Tahsina Khatun, Phazha Bothongo, Dylan M Williams, Jasmine Khouja, Emma L Anderson

## Abstract

**Introduction:** The long-term nicotine effects on neurocognitive outcomes remain unclear. This knowledge gap is critical given rising nicotine product use in the UK, and the global dementia burden. Polygenic risk score (PRS) analyses allow us to explore the direct effects of nicotine on neurocognitive outcomes in a way that mitigates bias by confounding and reverse causation, which current research has not been able to achieve so far. We utilised PRSs for nicotine metabolite ratio (NMR) and cigarettes smoked per day (CPD) to investigate the direct effects of nicotine on a range of neurocognitive outcomes.

**Methods:** Using 334,757 participants from the UK Biobank cohort study, we tested whether an NMR PRS affects the risk of six major neurocognitive outcomes: all-cause dementia (ACD), Alzheimer’s disease (AD), all-cause Parkinson’s disease (ACP), vascular dementia (VAD), total white matter hyperintensity volume (WMHv), and fluid intelligence score (FI).

**Results:** Increased nicotine metabolism (indicating increased nicotine clearance from the body and therefore reduced nicotine exposure per cigarette smoked) was associated with decreased risk of AD in multivariable analyses with CPD adjusted for (OR = 0.95, 95% CI 0.91 to 1.00) (i.e. increased nicotine exposure increases AD risk). Additionally, smoking heaviness adversely affected WMHv in multivariable analyses adjusting for NMR (β = 0.04, 95% CI -0.005 to 0.09). Our findings for nicotine and smoking heaviness in relation to our other neurocognitive outcomes indicate small effect sizes and confidence intervals cross the null.

**Conclusion:** Our findings demonstrate differential effects of nicotine on AD risk and smoking intensity on white matter integrity, requiring triangulation with complementary methodological approaches to better inform potential long-term risks of nicotine intake on neurocognitive outcomes.

## Introduction

Cigarette smoking is a well-established risk factor for neurocognitive outcomes such as Alzheimer’s disease^1,2^, vascular dementia^3^, and cerebral small vessel disease^4^, but some studies have reported a protective effect of smoking on Parkinson’s disease (PD)^5^ and overall cognition^6,7^. These neuroprotective effects are thought to be mediated by nicotine, tobacco’s primary psychoactive component. Nicotine use has increased in the UK^8^ in recent years due to the popularity of nicotine-based products, such as e-cigarettes (as a substitute for combustible cigarette smoking) and nicotine pouches^9^. However, the risks of long-term exposure to nicotine regarding neurocognitive outcomes remain unclear. Given that dementia represents a major global health burden^10^, identifying modifiable risk factors like nicotine is crucial for reducing the overall prevalence and impact of the disease.

Research on tobacco and nicotine’s neurocognitive effects has produced conflicting results^2,11–19^, potentially due to methodological limitations. Most of the current literature is observational and cannot establish causation. This evidence is also susceptible to limitations such as confounding bias distorting associations, or reverse causation, by which the early stages of the disease process or the diagnosis itself may influence smoking behaviours^20,21^. Survival bias may also potentially obscure these findings due to early fatal cardiovascular events or cancer linked to smoking^22–24^, which prevent diagnosis with neurocognitive outcomes later in life. Additionally, some of the evidence supporting a protective effect of specifically nicotine on neurocognitive outcomes is derived from experimental animal studies^11,14,15^, which are limited by their inability to produce translatable findings to humans.

Accordingly, evidence from studies addressing these limitations is needed. Achieving this by conducting a randomised controlled trial (RCT)^21,25–27^ would be unethical (as it would require nicotine-naive participants to be exposed to a potential harm for a sustained period), but genetic epidemiological techniques can be used as an alternative. One example is through the use of polygenic risk scores (PRS). Genome-wide association studies (GWAS) identify single nucleotide polymorphisms (SNPs) that are associated with a complex trait or disease^28^ (such as nicotine intake). The effects of each of these SNPs, generally, range between low to modest effects, but they can be summed to form a PRS for the trait or disease of interest. In this way, PRSs can be considered as an estimate of an individual’s genetic predisposition to a trait or disease^29^, and can be used as a proxy for exposure to a phenotype/trait. By using SNPs, PRS analyses exploit the unique properties of genetic variants; they are randomly inherited at conception^29^ (comparable to the random allocation of participants to exposure groups in RCTs), making confounding by environmental and socioeconomic factors highly unlikely as genetic variants cannot be influenced by confounding factors occurring later in life. Furthermore, because this random distribution of SNPs takes place before any outcomes occur (paralleling the temporal structure of RCTs, where the exposure is assigned first and the outcome is assessed after), reverse causation is avoided, making the investigation of causal relationships between exposure and downstream outcomes more plausible. In this way, PRSs can be used to explore causal relationships and mitigate common limitations of observational research in a way that resembles the framework of RCTs.

By combining SNPs *(SNP dosage*coefficient)* that have been identified in large-scale GWAS to be associated with nicotine metabolite ratio (NMR) (i.e., the rate at which an individual metabolises nicotine^30^) to construct an NMR PRS, it is possible to isolate nicotine from non-nicotinic constituents of tobacco smoke^21^. This approach allows for an in-depth investigation into nicotine’s relationship with neurocognitive outcomes in a way that previous research has not been able to achieve so far by overcoming key limitations.

Focusing solely on NMR as the exposure, however, may provide ambiguous results; if an individual has a high NMR, this is usually interpreted as a faster clearance of nicotine from their system, and thus a lower exposure to nicotine. This may also mean that these individuals are likely to smoke more^21^, resulting in a higher exposure to nicotine over time. Therefore, it is unclear as to whether a higher NMR alone translates to more or less nicotine exposure. To account for this, we can control for smoking heaviness in a multivariable model; an approach which has been employed in previously published literature^21^. Through an adaptation of this approach, the direct impact of nicotine (using the NMR PRS) on neurocognitive outcomes can be assessed while simultaneously fixing the amount of cigarettes smoked (using a PRS for cigarettes smoked per day [CPD]). By doing so, an increase in NMR (per cigarette smoked) can be interpreted as increased nicotine clearance from the body, i.e., reduced exposure to nicotine.

In this study, we aimed to use genetic instruments (PRSs) for NMR and CPD, to investigate the direct effects of nicotine on a range of neurocognitive outcomes (all-cause dementia [ACD], Alzheimer’s disease [AD], all-cause Parkinsonism [ACP], vascular dementia [VAD], total white matter hyperintensity volume [WMHv], and fluid intelligence [FI] score) using a genetic epidemiological approach designed to minimise common sources of bias in a large population sample.

## Methods

### Study participants

Participants were from UK Biobank (UKB), a cohort of 502,650 UK individuals. Details of the recruitment process and eligibility criteria for UKB have been described previously^31,32^. Detailed information on the health and lifestyle of participants was obtained from regular assessment centre visits, completion of questionnaires, health-related records, and blood, urine, and saliva samples^33^. Data details are available at biobank.ndph.ox.ac.uk/showcase/. Details of the genotyping process of UKB participants have also been described previously^34–36^.

Participants with available genotype and phenotype data were eligible for inclusion in our analyses. We excluded: sex mismatches, aneuploidies, heterozygosity outliers, withdrawn consent, and third-degree relatives^35^. We restricted to genetically-defined White British participants for homogeneity, as the NMR GWAS used for this study was European-ancestry specific. Figure 1 displays a flowchart outlining the steps taken to define the final analytical sample.

**Fig 1.**
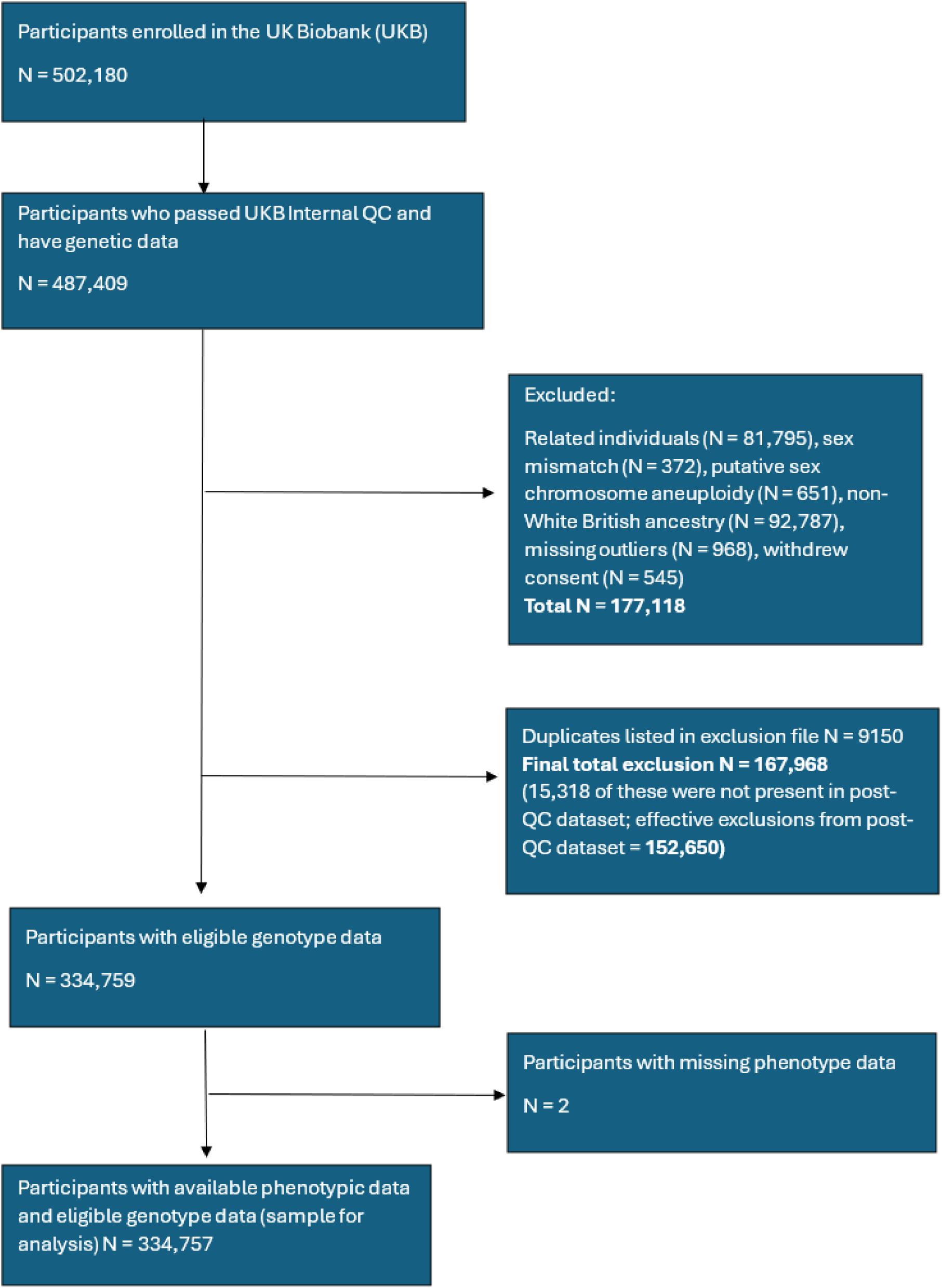
Flowchart illustrating how the analytical sample size was defined from the initial UKB sample. Abbreviations: QC, quality control

### GWAS data

To derive the NMR PRS, summary statistics were obtained from the largest NMR GWAS meta-analysis to date, in smokers of European descent, conducted by Buchwald and colleagues^37^. To derive the CPD PRS, summary statistics were retrieved from a GWAS meta-analysis of smoking and alcohol phenotypes (including CPD) from individuals of European ancestry only, performed by the GWAS and Sequencing Consortium of Alcohol and Nicotine use (GSCAN)^38^. To avoid sample overlap, we used a version of the GWAS data for CPD where UK Biobank data were removed.

### Smoking phenotypes

As part of the UKB participant questionnaire, participants are asked questions surrounding current and previous tobacco smoking (field IDs of all UKB variables are listed in the Supplementary Materials). UKB categorised participants as Never/Previous/Current smokers. We pooled Previous and Current into ’Ever’ smokers; Never remained unchanged. Participants who answered ‘Prefer not to answer’ were classed as missing. Our main analyses focused on the subgroup of participants who were classed as ‘ever’ smokers, because the NMR and CPD GWAS were conducted among smokers only, thus, our constructed PRSs are only valid among individuals who smoke. For this reason, we expect to see no effect of the NMR or CPD PRSs among people who do not smoke, therefore, the subgroup of participants belonging to ‘never’ smokers were used for negative control analyses.

### Neurocognitive outcomes

The following outcomes were investigated: ACD, AD, ACP, VAD, WMHv, and FI. These outcomes were chosen as previous evidence suggests they are influenced by smoking activity and behaviour^2–5,39,40^.

ACD, AD, ACP and VAD data were retrieved from UKB’s algorithmically-defined outcomes, described in the Supplementary Materials and by UKB^41^. For PD in particular, UKB’s algorithm provides measures for two variables: PD and all-cause Parkinsonism. We used all-cause Parkinsonism for broader coverage and larger sample size purposes.

WMHv (a neuroimaging biomarker of cerebral small vessel disease) neuroimaging commenced in 2014. If participants were missing data from the initial baseline neuroimaging assessment, values were replaced with WMHv measured at the follow-up assessment where available. WMHv measures were acquired using T1 and T2_FLAIR MRI sequences, in line with UKB’s brain imaging pipeline, described previously^42^. Due to the average age of UKB participants and the healthy selection into UKB^43^, WMHv measures within the sample were skewed (with most participants presenting low volumes of white matter hyperintensities). Therefore, the WMHv variable was log-transformed to normalise the distribution of the data^44^, and then standardised to ensure all continuous variables within our analyses (FI and WMHv) were interpreted on the same standard deviation scale.

For FI (a cognitive battery consisting of 13 questions, designed to measure cognitive function across several aspects), we used baseline scores from the initial assessment if available. Missing values were replaced with available FI scores measured online at the follow-up assessment approximately 4-7 years later. Full details of the FI score can be found online^45^ and in the Supplementary Materials. The FI variable was also standardised here.

### Statistical analyses

All analyses were performed in RStudio 4.4.1.

### Generating polygenic risk scores

Polygenic risk scores were derived in accordance with the UKB PRS pipeline described previously^35^. We obtained summary-level genetic data from the NMR and CPD GWAS described above. Briefly, for each participant, the SNP dosage for all independent, genome-wide significant variants was multiplied by their respective weight taken from the relevant GWAS, and the results were summed across all variants. PRSs need to be weighted in order to reflect the relative magnitude of effect of each SNP on the trait of interest^29^. We included only biallelic SNPs, excluding 10kb around *APOE*. *APOE* exclusion prevented confounding by ε4 allele’s effect on dementia and neurocognitive outcomes overall^46–49^. A list of all SNPs used to derive the NMR (n=6 SNPs) and CPD (n=55 SNPs) PRSs, at the genome-wide significant threshold, is provided in the Supplementary Materials.

### Regression models

Univariable analyses were initially conducted to assess the associations between the NMR and CPD PRS individually with the outcomes. Models adjusted for age, sex, batch, centre, and 10 genetic principal components (PCs). We then fitted multivariable regression models including both the NMR and CPD PRSs as exposures of interest, to estimate the effect of each PRS on the outcomes, while controlling for the other PRS. The same covariate adjustments were made in the multivariable models.

Both univariable and multivariable analyses were conducted among ever smokers. Linear and logistic regression models were fitted for continuous and binary outcomes, respectively.

In multivariable analyses, for continuous outcomes (WMHv and FI) with CPD PRS adjusted for as a proxy for smoking heaviness, we interpret the direct effects of nicotine by taking the inverse of the NMR beta i.e., a positive beta for NMR PRS in the multivariable analyses is interpreted as a reduction in systemic nicotine exposure per cigarette smoked, and a negative beta for the NMR PRS in the multivariable analyses is interpreted as an increase in nicotine exposure per cigarette smoked. For binary outcomes (ACD, AD, ACP and VAD) with CPD adjusted for, odds ratios (OR) are reported, whereby an NMR PRS OR above 1 indicates that with a higher NMR (i.e., lower nicotine exposure), there are increased odds of the outcome. An NMR PRS OR below 1 indicates that with a higher NMR (i.e., lower nicotine exposure), there are decreased odds of the outcome. The results are interpreted here based on the magnitude of associations and their respective confidence intervals, rather than highlighting results for which the *p*-value is below an arbitrary threshold of <0.05^50^.

### Instrument validation analysis

Nicotine increases heart rate^21^, and the constructed CPD PRS should be highly predictive of the number of cigarettes smoked daily by an individual, thus, we examined these known associations in an instrument validation analysis to assess the validity of our PRSs. The NMR instrument validation analysis was restricted to current smokers only, as the effect of nicotine on heart rate is acute^51^ and therefore is not applicable to ‘previous’ smokers, as they are no longer exposed to nicotine. CPD instrument validation analyses were conducted in ever smokers (as the PRS is valid in smokers only, and to remain consistent with the main analyses). For CPD, two phenotypic measures are available in UKB – the number of cigarettes currently smoked per day (among current smokers) and the number of cigarettes previously smoked per day (among former smokers), so the CPD PRS was tested with both of these phenotypes individually, as well as with a third phenotype: the number of cigarettes ever smoked per day, which was derived by pooling data from the former two phenotypes. In all instrument validation analyses, the same covariates were adjusted for (age at recruitment, sex, genotype measurement batch, UKB assessment centre, and the first 10 genetic PCs). In the NMR instrument validation analyses, models were additionally adjusted for CPD PRS again for clearer interpretation of the nicotine results.

### Negative control analysis

Univariable and multivariable analyses were repeated in never smokers, with never smokers serving as a negative control group, as both PRSs should only have an effect in smokers.

### Sensitivity analyses

All analyses were repeated using an alternative CPD PRS, derived from summary statistics from individuals of European ancestry reported by GSCAN in an updated GWAS, conducted by Saunders and colleagues^52^. As standard errors were consistently higher in both univariable and multivariable analyses using this alternative CPD PRS (compared to the primary analyses), therefore giving rise to results with less precision, we treated this analysis as a sensitivity analysis, with the aim of evaluating the robustness of our results.

## Results

### Sample description

A total of 334,757 participants had available genetic and phenotype data and were eligible for inclusion in analyses. Table 1 presents descriptive statistics by smoking status.

**Table 1:** Characteristics of participants in the analytical sample. All participants were of European ancestry.

|  | UK Biobank participants |  |
| --- | --- | --- |
|  | Ever smokers | Never smokers |
| N | 150,736 | 182,847 |
| Sex = Male (%) | 78,452 (52%) | 75,648 (41%) |
| Mean age at recruitment (SD) | 57.7 (7.8) | 56.2 (8.1) |
| ACD diagnosis n (% of sample) | 3591 (2%) | 3081 (2%) |
| AD diagnosis n (% of sample) | 1550 (1%) | 1423 (<1%) |
| VAD diagnosis n (% of sample) | 847 (<1%) | 629 (<1%) |
| ACP diagnosis n (% of sample) | 1532 (1%) | 1768 (1%) |
| Mean log standardised WMHv (SD) | 0.12 (1.0) | -0.08 (1.0) |
| Mean standardised FI score (SD) | -0.04 (1.0) | 0.03 (1.0) |

Abbreviations: ACD, all-cause dementia; ACP, all-cause Parkinsonism; AD, Alzheimer's disease; FI, fluid intelligence; SD, standard deviation; VAD, vascular dementia; WMHv, white matter hyperintensity volume.

### Instrument validation analysis (heart rate and CPD phenotypes)

There was uncertain evidence of an effect of NMR on heart rate in the multivariable analysis (β = -0.10, 95% CI -0.23 to 0.04) in current smokers. Although the confidence intervals span the null, the direction of the effect aligns with previous knowledge of nicotine increasing heart rate^21^. Our instrument validation analyses for CPD consistently showed strong evidence supporting that our CPD PRS is associated with the number of cigarettes currently smoked per day (β = 2.74, 95% CI 2.41 to 3.08), the number of cigarettes previously smoked per day (β = 2.99, 95% CI 2.77 to 3.21), and the number of cigarettes ever smoked per day (β = 2.96, 95% CI 2.77 to 3.16), thus, validating the robustness of our constructed PRS for CPD.

### Primary analysis

The total and direct effects of the NMR and CPD PRSs on all outcomes: ACD, AD, ACP, VAD, WMHs, and FI, are presented in Figure 1 (ever smokers). Results relating to binary outcomes are reported as the odds of disease per standard deviation (SD) increase in the NMR or CPD PRS. Results for continuous outcomes are presented as standardised betas per SD increase in the NMR or CPD PRS. Full results are included in the Supplementary Materials.

**Figure 1:**
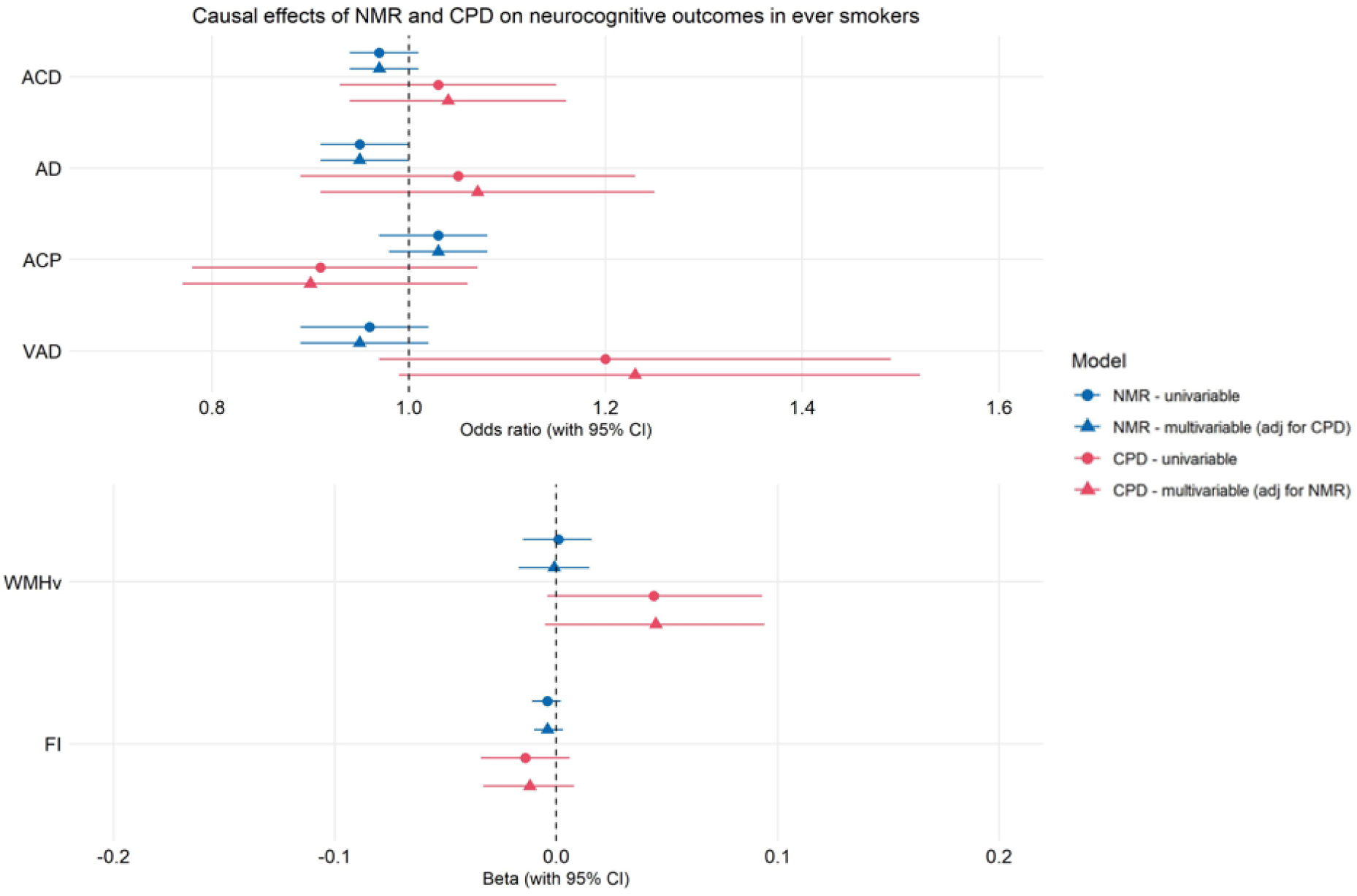
Forest plot displaying the causal effects of NMR PRS and CPD PRS on neurocognitive outcomes (all-cause dementia [ACD], Alzheimer’s disease [AD], all-cause Parkinsonism [ACP], vascular dementia [VAD], white matter hyperintensity volume [WMHv], and fluid intelligence [FI]) among ever smokers. Note: blue = NMR PRS estimates; red = CPD PRS estimates, circle = univariable models; triangle = multivariable models.

### Univariable effects – NMR PRS

There was weak evidence to suggest that increased NMR decreases the risk of AD (odds ratio [OR] 0.95, 95% confidence interval [CI] 0.91 to 1.00), ACD risk (OR = 0.97, 95% CI 0.94 to 1.01) and VAD risk (OR = 0.96, 95% CI 0.89 to 1.02), although confidence intervals cross the null (for ACD and VAD). The results for the NMR PRS on our other outcomes were as follows: ACP risk (OR = 1.03, 95% CI 0.97 to 1.08), WMHv (beta [β] = 0.001, 95% CI -0.02 to 0.02), and FI (β = -0.004, 95% CI -0.01 to 0.002), among ever smokers, with confidence intervals crossing the null.

### Univariable effects – CPD PRS

The univariable results provide weak evidence that increased CPD PRS increases WMHv (β = 0.04, 95% CI -0.004 to 0.09), and weaker evidence that increased CPD PRS increases VAD risk (OR = 1.20, 95% CI 0.97 to 1.49), but this effect is not clear, with confidence intervals just crossing the null. Additionally, for the CPD PRS, the results were: ACD risk (OR = 1.03, 95% CI 0.93 to 1.15), AD risk (OR = 1.05, 95% CI 0.89 to 1.23), ACP risk (OR = 0.91, 95% CI 0.78 to 1.07), and FI (β = -0.01, 95% CI -0.03 to 0.01) among ever smokers in univariable analyses, with confidence intervals crossing the null.

### Multivariable effects – NMR PRS (with CPD PRS adjusted for)

Multivariable NMR results (CPD-adjusted) supported univariable findings, whereby increased NMR PRS (indicative of decreased nicotine exposure) showed weak evidence of lowering AD risk (OR = 0.95, 95% CI 0.91 to 1.00), while there was weaker evidence of an effect for the NMR PRS on ACD risk (OR = 0.97, 95% CI 0.94 to 1.01) and VAD risk (OR = 0.95, 95% CI 0.89 to 1.01), with confidence intervals overlapping the null. For our other outcomes, the multivariable results for the NMR PRS were: ACP risk (OR = 1.03, 95% CI 0.98 to 1.08), WMHv (β = -0.001, 95% CI -0.02 to 0.01), and FI (β = -0.004, 95% CI -0.01 to 0.003), among ever smokers, with confidence intervals crossing the null.

### Multivariable effects – CPD PRS (with NMR PRS adjusted for)

Similarly to univariable analyses, in multivariable analyses (with NMR adjusted for), there was weak evidence of a causal effect of CPD PRS on WMHv (β = 0.04, 95% CI -0.005 to 0.09) and VAD risk (OR = 1.23, 95% CI 0.99 to 1.52), with confidence intervals crossing the null by a small amount. The relationship between the CPD PRS and our other outcomes were as follows: ACD risk (OR = 1.04, 95% CI 0.94 to 1.16), AD risk (OR = 1.07, 95% CI 0.91 to 1.25), ACP risk (OR = 0.90, 95% CI 0.77 to 1.06) and FI (β = -0.01, 95% CI -0.03 to 0.01) among ever smokers, with confidence intervals crossing the null here.

### Negative control analysis (never smokers)

In never smokers (negative controls), analyses showed no evidence of an effect of NMR or CPD PRS on any of our outcomes, except for WMHv. There was weak evidence suggesting that increased NMR increases WMHv in univariable analyses (β = 0.01, 95% CI 0.001 to 0.03) and in multivariable analyses with CPD adjusted for (β = 0.01, 95% CI 0.001 to 0.03) (indicative of decreased nicotine exposure when accounting for CPD).

### Instrument strength

Conditional F-statistics represent the strength of the genetic instrument in measuring one exposure while accounting for other exposures included in the calculation^53^. A conditional F-statistic greater than 10 indicates a strong genetic instrument. As UKB does not hold phenotypic data for nicotine intake or NMR, conditional F-statistics could not be calculated for the NMR and CPD PRSs, however, conditional F-statistics for the same set of NMR and CPD SNPs have been previously published^21^ (F=49.08 for NMR and 34.17 for CPD), demonstrating that the SNPs used here in our analyses should be strong genetic instruments for investigating the effects of NMR and CPD in all analyses.

### Sensitivity analyses

Results obtained from analyses using the alternative CPD PRS are presented in the Supplementary Materials. Generally, these results were consistent with the main results (supporting the robustness of our findings), with additional insights. In multivariable analyses adjusting for the alternative CPD PRS, there was slightly more evidence to suggest NMR increases ACD risk (OR = 0.97, 95% CI 0.93 to 1.00) and VAD risk (OR = 0.93, 95% CI 0.86 to 1.00) in ever smokers than we observed in the main analysis, as confidence intervals did not cross the null in the sensitivity analysis, but did cross the null in the main analysis. Furthermore, when using this alternative CPD PRS, there was evidence to suggest increased CPD PRS reduces FI scores (i.e., worse cognition) in both univariable (β = -0.04, 95% CI -0.07 to -0.01) and multivariable analyses (β = -0.03, 95% CI -0.07 to -0.0009) among ever smokers.

However, standard errors were consistently higher in both univariable and multivariable analyses utilising this alternative CPD PRS (in comparison to analyses using the primary CPD PRS), therefore giving rise to results with less precision.

## Discussion

To our knowledge, this is the first study disentangling the effects of nicotine from tobacco smoke on multiple neurocognitive outcomes using PRSs. For ease of interpretation, we discuss AD, ACD, and VAD results in terms of increased nicotine exposure, in contrast with the results section, where the effects are described in terms of decreased nicotine exposure. Overall, our results suggest that, among ever smokers, increased nicotine exposure (per cigarette smoked) increases risk of AD. The effects of nicotine on our other neurocognitive outcomes (ACD, ACP, VAD, WMHs, and FI) in both univariable and multivariable models are unclear as confidence intervals overlap the null.

Our findings provide weak evidence that is in line with previous evidence suggesting that nicotine may increase AD risk in various ways. Chronic nicotine exposure induces cerebral oxidative stress^2,54^, which in turn induces production of biomarkers that are central to AD pathology; namely, aggregated amyloid-beta (aβ) (through means of promoting increased β-secretase cleavage of the amyloid precursor protein)^55,56^, as well as general neurodegeneration^57^. Nicotine also disrupts blood-brain barrier integrity and function^58–61^, leading to impaired clearance of aβ from the brain and thus, facilitating the buildup of aβ in the brain^62^. Furthermore, nicotine is associated with insomnia and disrupted sleeping patterns^63,64^, which, in turn, impairs the glymphatic system from clearing toxic waste products (such as aβ) from the brain, again promoting cerebral aβ accumulation^65^. Through these mechanisms, nicotine can favour the onset of AD. However, our effect sizes for AD are small (indicating a 5% increase in AD risk per SD reduction in the NMR PRS), so these results carry a degree of uncertainty.

Additionally, we observed suggestive evidence that nicotine increases ACD and VAD risk, although effect sizes were small and confidence intervals just crossed the null. Studies investigating nicotine’s role in ACD and VAD are scarce, although some studies show chronic nicotine exposure promotes oxidative stress and endothelial dysfunction, leading to reduced blood flow to the brain^66^ – a primary mechanism that is central to the onset of vascular dementia^67,68^. There is also a lack of research on the role of nicotine on WMHs, therefore, our results present novel findings for ACD, VAD, and WMHv. Additional studies are required to validate our results and explore the effect of nicotine on these neurocognitive outcomes further.

In contrast, a large body of research over the last few decades suggests that nicotine is protective against PD^14,69^. Our findings do not provide strong support for this inverse relationship. Although our effect size is directionally consistent with a beneficial effect of nicotine on ACP, the confidence intervals cross the null, so there may not be a true effect. Our effect estimate is also small (indicating a 3% reduction in PD risk per SD reduction in the NMR PRS), and so any advantageous effect of nicotine on ACP risk may be minimal. The presence of selection bias into UKB^70^ may explain why we do not see a strong effect; as the average age of recruitment into UKB in our analytical sample was ∼56 years and symptoms of PD can occur earlier than this^71^, which may have impacted enrolment into the study.

Where previous studies suggest nicotine enhances several cognitive domains^72^ via binding to its receptors in cognitive-rich regions of the brain^6^, we did not find clear evidence of an effect of nicotine on FI. Many of the studies which demonstrate that nicotine improves cognition involve short-term administration of nicotine, and therefore any beneficial impact nicotine has on cognition is likely to be acute and temporary, and does not reflect lifelong exposure to nicotine. Chronic nicotine exposure leads to desensitisation of their receptors^73^, which in turn makes the receptors less responsive to nicotine upon binding, attenuating the cognitive-enhancing effects of nicotine. Here, we used PRSs that act as genetic instruments capturing lifelong exposure to nicotine and therefore are a better representation of real-life smoking and nicotine consumption.

Considering smoking heaviness more broadly, we observed weak evidence that increased smoking heaviness detrimentally impacts WMHv (and risk of VAD, to a less clear extent) among ever smokers, in agreement with previous research^4,40,74^. We present evidence suggesting that smoking heaviness influences ACD, AD, ACP, and FI to some extent, but the true effects are unclear here due to confidence intervals crossing the null more markedly.

For PRS analyses to yield valid causal insights, three instrumental variable (IV) assumptions must be met: IV1) the genetic instrument (the PRSs) is robustly associated with the exposure of interest, IV2) there is no confounding of the genetic instrument-outcome relationship, and IV3) the genetic instrument affects the outcome only via the exposure (i.e. there are no pleiotropic pathways present)^75^. We have made efforts to assess all three assumptions. Although formal methods have not been developed to identify pleiotropy in PRS analyses^29^, we attempted to minimise pleiotropy by using smoking heaviness as an exposure, which is considered a less pleiotropic measure compared to other smoking measures (such as smoking initiation)^76^. We also informally tested for potential pleiotropy by conducting analyses between the PRSs and a range of measures that could lie on the pathway between the PRS and our neurocognitive outcomes. Associations were observed for the NMR PRS with body mass index (BMI) in univariable and multivariable analyses. Although this may indicate the presence of pleiotropic genetic variants within our NMR PRS, these associations may be driven by other mechanisms, such as confounding bias or selection bias, which could induce spurious associations between the PRS and BMI, making it difficult to determine if the IV3 assumption has been met here.

### Strengths and limitations

Using cohort data from UKB enabled the use of deeply phenotyped data across multiple aspects, including biological, clinical, and demographic factors, enabling us to carry out an in-depth investigation into the complex genetic relationship between nicotine and smoking heaviness and their impact on neurocognitive outcomes. A major strength of our study is that the use of genetic scores as proxies for smoking and nicotinic behaviour reduces the impact of limitations that often weaken traditional observational research findings. Furthermore, results from our instrument validation analyses are generally in line with what was expected. These results, taken together with the high conditional F-statistics which have been published elsewhere for the same set of genetic variants^21^, indicate our PRSs of NMR and CPD are robust genetic predictors of nicotine intake and smoking activity.

There are several limitations to this study. UKB recruited adults aged 40-69 who were mostly healthy at the time of recruitment^43,70^, reducing the likelihood of inclusion of those with early-onset symptoms or diagnoses. There are also a limited number of cases for each of the outcomes, therefore we may have had limited power to detect an effect. Our findings also cannot be generalised to other populations, as the PRSs were derived using summary statistics from European participants, and we restricted the analytical sample to White participants of European ancestry only. There is a disparity in neurocognitive outcomes between different racial and ethnic groups, with an increased burden in minority groups^77^. Nicotine metabolite rates also differ between different ethnic groups – generally, non-White participants metabolise nicotine much slower compared to White participants^78,79^. Incorporating other diverse populations into future studies will lead to inclusive results which can be applied universally and may provide a more holistic overview of nicotine’s influence on neurocognitive outcomes. However, such an analysis requires a large, multi-ancestry/multi-ethnic GWAS of NMR and of some of our outcome data, which are not currently available. Additionally, the use of self-reported data on smoking status and the number of cigarettes smoked per day can give rise to unreliable smoking data due to differences in smoking topography^80,81^. Furthermore, stratifying the sample by smoking status reduces power and potentially introduces collider bias^21,76^. Negative control analyses should have shown no effects of NMR and CPD with all outcomes among never smokers, yet we observed a protective effect of NMR on WMHv. This result may be due to chance, as multiple associations were tested without adjusting for correction. This can also indicate potential bias from various sources, including (but not limited to) horizontal pleiotropy, residual population stratification, dynastic effects, assortative mating, and measurement error in the ‘ever’ vs ‘never’ smoking status, although efforts were made to minimise these sources of bias.

### Future work

Replication in other cohorts is required to validate our findings. The use of alternative cohorts – particularly case-control cohorts and more ethnically diverse cohorts – will strengthen the generalisability of the results. Other indicators of cognitive function, such as fractional anisotropy^82,83^ and mean diffusivity^84^, should be incorporated to further explore nicotine’s impact on more nuanced measures of cognition. The impact of nicotine on other dementia-causing neurocognitive outcomes not assessed here should also be explored, including, but not limited to, Huntington’s disease and dementia with Lewy bodies, given their ties to smoking^85,86^. We were unable to conduct Mendelian randomisation (MR) analyses here due to a lack of access to smoking-stratified GWAS data which are required for two-sample MR^87^, so MR analyses are warranted to validate our results and strengthen causal inference. Although traditionally, MR is a widely used genetic method used to make causal inferences^88^, PRS analyses also share many conceptual and practical similarities with MR, particularly in aetiological epidemiology, and findings from PRS analyses are often comparable to those from MR analyses (in analyses where heterogeneity is low and the PRS is scaled to the exposure)^29^.

## Conclusion

In conclusion, increased nicotine intake (when accounting for smoking heaviness) appears to increase the risk of AD among ever smokers, and the effects of nicotine on our other neurocognitive outcomes are small, with confidence intervals crossing the null. The discrepancy between our results and existing results emphasises the value of using genetically informed methods to clarify causal relationships. Insights from this study, together with results from future research, can be triangulated to contribute to more informative education on the potential long-term risks surrounding e-cigarette use and nicotine intake.

## Data availability

All GWAS data used in the study are publicly available. NMR GWAS data can be accessed at https://pmc.ncbi.nlm.nih.gov/articles/PMC7483250/. Summary statistics used to construct the CPD PRS applied in main analyses can be found at https://conservancy.umn.edu/items/ca7ed549-636b-41c0-ae79-97c57e266417. Summary statistics used to construct the CPD PRS applied in sensitivity analyses can be found at https://conservancy.umn.edu/items/91f6a003-6af2-4809-9785-53dc579dc788.

## Ethics statement

All participants provided written informed consent for participation in UK Biobank. Approval to use UK Biobank data for this study was approved by UK Biobank (project 123335). UK Biobank attained ethical approval by the North West Multi-centre Research Ethics Committee as a Research Tissue Bank approval^89^ (reference 11/NW/0382), therefore all projects approved by UKB do not require individual ethical clearance.

## Bibliography

1. Peters R, Poulter R, Warner J, Beckett N, Burch L, Bulpitt C. Smoking, dementia and cognitive decline in the elderly, a systematic review. BMC Geriatr. 2008;8:36. doi:10.1186/1471-2318-8-36

2. Durazzo TC, Mattsson N, Weiner MW. Smoking and increased Alzheimer’s disease risk: A review of potential mechanisms. Alzheimer’s & dementia : the journal of the Alzheimer’s Association. 2014;10(3 0):S122. doi:10.1016/j.jalz.2014.04.009

3. Rusanen M, Kivipelto M, Quesenberry CP, Zhou J, Whitmer RA. Heavy smoking in midlife and long-term risk of Alzheimer disease and vascular dementia. Arch Intern Med. 2011;171(4):333–339. doi:10.1001/archinternmed.2010.393

4. Power MC, Deal JA, Sharrett AR, et al. Smoking and white matter hyperintensity progression. Neurology. 2015;84(8):841–848. doi:10.1212/WNL.0000000000001283

5. Hong SW, Page R, Truman P. Smoking, coffee intake, and Parkinson’s disease: Potential protective mechanisms and components. NeuroToxicology. 2025;106:48–63. doi:10.1016/j.neuro.2024.12.003

6. Wang Q, Du W, Wang H, et al. Nicotine’s effect on cognition, a friend or foe? Progress in Neuro-Psychopharmacology and Biological Psychiatry. 2023;124:110723. doi:10.1016/j.pnpbp.2023.110723

7. Momtaz YA, Ibrahim R, Hamid TA, Chai ST. Smoking and Cognitive Impairment Among Older Persons in Malaysia. American Journal of Alzheimer’s Disease & Other Dementias®. Published online September 26, 2014. doi:10.1177/1533317514552318

8. Tattan-Birch H, Brown J, Shahab L, Beard E, Jackson SE. Trends in vaping and smoking following the rise of disposable e-cigarettes: a repeat cross-sectional study in England between 2016 and 2023. Lancet Reg Health Eur. 2024;42:100924. doi:10.1016/j.lanepe.2024.100924

9. Brose L, Bunce L, Cheeseman H. Prevalence of Nicotine Pouch Use Among Youth and Adults in Great Britain—Analysis of Cross-Sectional, Nationally Representative Surveys. Nicotine Tob Res. doi:10.1093/ntr/ntae295

10. Steinmetz JD, Seeher KM, Schiess N, et al. Global, regional, and national burden of disorders affecting the nervous system, 1990–2021: a systematic analysis for the Global Burden of Disease Study 2021. The Lancet Neurology. 2024;23(4):344–381. doi:10.1016/S1474-4422(24)00038-3

11. Xue MQ, Liu XX, Zhang YL, Gao FG. Nicotine exerts neuroprotective effects against β-amyloid-induced neurotoxicity in SH-SY5Y cells through the Erk1/2-p38-JNK-dependent signaling pathway. International Journal of Molecular Medicine. 2014;33(4):925–933. doi:10.3892/ijmm.2014.1632

12. Lin X, Li Q, Pu M, Dong H, Zhang Q. Significance of nicotine and nicotinic acetylcholine receptors in Parkinson’s disease. Front Aging Neurosci. 2025;17. doi:10.3389/fnagi.2025.1535310

13. Barreto GE, Iarkov A, Moran VE. Beneficial effects of nicotine, cotinine and its metabolites as potential agents for Parkinson’s disease. Front Aging Neurosci. 2015;6:340. doi:10.3389/fnagi.2014.00340

14. Parain K, Hapdey C, Rousselet E, Marchand V, Dumery B, Hirsch EC. Cigarette smoke and nicotine protect dopaminergic neurons against the 1-methyl-4-phenyl-1,2,3,6-tetrahydropyridine Parkinsonian toxin. Brain Res. 2003;984(1-2):224–232. doi:10.1016/s0006-8993(03)03195-0

15. Srivareerat M, Tran TT, Salim S, Aleisa AM, Alkadhi KA. Chronic nicotine restores normal Aβ levels and prevents short-term memory and E-LTP impairment in Aβ rat model of Alzheimer’s disease. Neurobiology of Aging. 2011;32(5):834–844. doi:10.1016/j.neurobiolaging.2009.04.015

16. Wallin C, Sholts SB, Österlund N, et al. Alzheimer’s disease and cigarette smoke components: effects of nicotine, PAHs, and Cd(II), Cr(III), Pb(II), Pb(IV) ions on amyloid-β peptide aggregation. Sci Rep. 2017;7(1):14423. doi:10.1038/s41598-017-13759-5

17. Gil SM, Metherate R. Enhanced Sensory–Cognitive Processing by Activation of Nicotinic Acetylcholine Receptors. Nicotine Tob Res. 2018;21(3):377–382. doi:10.1093/ntr/nty134

18. Oddo S, Caccamo A, Green KN, et al. Chronic nicotine administration exacerbates tau pathology in a transgenic model of Alzheimer’s disease. Proc Natl Acad Sci U S A. 2005;102(8):3046–3051. doi:10.1073/pnas.0408500102

19. Deng J, Shen C, Wang YJ, et al. Nicotine exacerbates tau phosphorylation and cognitive impairment induced by amyloid-beta 25-35 in rats. Eur J Pharmacol. 2010;637(1-3):83–88. doi:10.1016/j.ejphar.2010.03.029

20. Ritz B, Lee PC, Lassen CF, Arah OA. Parkinson disease and smoking revisited. Neurology. 2014;83(16):1396-1402. doi:10.1212/WNL.0000000000000879

21. Khouja JN, Sanderson E, Wootton RE, et al. Estimating the health impact of nicotine exposure by dissecting the effects of nicotine versus non-nicotine constituents of tobacco smoke: A multivariable Mendelian randomisation study. PLOS Genetics. 2024;20(2):e1011157. doi:10.1371/journal.pgen.1011157

22. Thomson B, Emberson J, Lacey B, Peto R, Woodward M, Lewington S. Childhood Smoking, Adult Cessation, and Cardiovascular Mortality: Prospective Study of 390 000 US Adults. J Am Heart Assoc. 2020;9(21):e018431. doi:10.1161/JAHA.120.018431

23. Khan SS, Ning H, Sinha A, et al. Cigarette Smoking and Competing Risks for Fatal and Nonfatal Cardiovascular Disease Subtypes Across the Life Course. J Am Heart Assoc. 2021;10(23):e021751. doi:10.1161/JAHA.121.021751

24. Dragani TA, Muley T, Schneider MA, et al. Lung Adenocarcinoma Diagnosed at a Younger Age Is Associated with Advanced Stage, Female Sex, and Ever-Smoker Status, in Patients Treated with Lung Resection. Cancers (Basel). 2023;15(8):2395. doi:10.3390/cancers15082395

25. Braga LH, Farrokhyar F, Dönmez Mİ, et al. Randomized controlled trials – The what, when, how and why. Journal of Pediatric Urology. 2025;21(2):397–404. doi:10.1016/j.jpurol.2024.11.021

26. Heindel P, Dieffenbach BV, Freeman NLB, McGinigle KL, Menard MT. Central concepts for randomized controlled trials and other emerging trial designs. Seminars in Vascular Surgery. 2022;35(4):424–430. doi:10.1053/j.semvascsurg.2022.10.004

27. Swerdlow DI, Michail M, Zacharias K. Chapter 8 - Role of Conventional Risk Factors in Genetic Susceptibility to Cardiovascular Diseases. In: Papageorgiou N, ed. Cardiovascular Diseases. Academic Press; 2016:159–176. doi:10.1016/B978-0-12-803312-8.00008-2

28. Korologou-Linden R, Anderson EL, Jones HJ, Davey Smith G, Howe LD, Stergiakouli E. Polygenic risk scores for Alzheimer’s disease, and academic achievement, cognitive and behavioural measures in children from the general population. Int J Epidemiol. 2019;48(6):1972–1980. doi:10.1093/ije/dyz080

29. Garfield V, Anderson EL. A brief comparison of polygenic risk scores and Mendelian randomisation. BMC Medical Genomics. 2024;17(1):10. doi:10.1186/s12920-023-01769-4

30. Siegel SD, Lerman C, Flitter A, Schnoll RA. The Use of the Nicotine Metabolite Ratio as a Biomarker to Personalize Smoking Cessation Treatment: Current Evidence and Future Directions. Cancer Prev Res (Phila). 2020;13(3):261–272. doi:10.1158/1940-6207.CAPR-19-0259

31. Sudlow C, Gallacher J, Allen N, et al. UK Biobank: An Open Access Resource for Identifying the Causes of a Wide Range of Complex Diseases of Middle and Old Age. PLoS Med. 2015;12(3):e1001779. doi:10.1371/journal.pmed.1001779

32. Hewitt J, Walters M, Padmanabhan S, Dawson J. Cohort profile of the UK Biobank: diagnosis and characteristics of cerebrovascular disease. BMJ Open. 2016;6(3):e009161. doi:10.1136/bmjopen-2015-009161

33. UK Biobank. Communications Guidelines. UK Biobank; 2025. Accessed November 27, 2025. https://community.ukbiobank.ac.uk/hc/en-gb/articles/24818044276765-Communications-guidelines

34. UK Biobank. Genetic data. Published online August 13, 2024. Accessed January 6, 2025. https://www.ukbiobank.ac.uk/enable-your-research/about-our-data/genetic-data

35. Collister JA, Liu X, Clifton L. Calculating Polygenic Risk Scores (PRS) in UK Biobank: A Practical Guide for Epidemiologists. Front Genet. 2022;13. doi:10.3389/fgene.2022.818574

36. UK Biobank. Genotyping of 500,000 UK Biobank Participants. UK Biobank; 2017. Accessed January 6, 2025. https://biobank.ctsu.ox.ac.uk/ukb/ukb/docs/ukb_dna_processing.pdf

37. Buchwald J, Chenoweth MJ, Palviainen T, et al. Genome-wide association meta-analysis of nicotine metabolism and cigarette consumption measures in smokers of European descent. Mol Psychiatry. 2021;26(6):2212–2223. doi:10.1038/s41380-020-0702-z

38. Liu M, Jiang Y, Wedow R, et al. Association studies of up to 1.2 million individuals yield new insights into the genetic etiology of tobacco and alcohol use. Nat Genet. 2019;51(2):237–244. doi:10.1038/s41588-018-0307-5

39. Juan D, Zhou DHD, Li J, Wang JYJ, Gao C, Chen M. A 2-year follow-up study of cigarette smoking and risk of dementia. Eur J Neurol. 2004;11(4):277–282. doi:10.1046/j.1468-1331.2003.00779.x

40. Anstey KJ, von Sanden C, Salim A, O’Kearney R. Smoking as a risk factor for dementia and cognitive decline: a meta-analysis of prospective studies. Am J Epidemiol. 2007;166(4):367–378. doi:10.1093/aje/kwm116

41. UK Biobank. UK Biobank Algorithmically-Defined Outcomes. UK Biobank; 2022. Accessed January 6, 2025. https://biobank.ndph.ox.ac.uk/showcase/showcase/docs/alg_outcome_main.pdf

42. UK Biobank. UK Biobank Brain Imaging Documentation. UK Biobank; 2024. Accessed January 6, 2025. https://biobank.ctsu.ox.ac.uk/crystal/crystal/docs/brain_mri.pdf

43. Fry A, Littlejohns TJ, Sudlow C, et al. Comparison of Sociodemographic and Health-Related Characteristics of UK Biobank Participants With Those of the General Population. Am J Epidemiol. 2017;186(9):1026–1034. doi:10.1093/aje/kwx246

44. Jochems ACC, Arteaga C, Chappell F, et al. Longitudinal Changes of White Matter Hyperintensities in Sporadic Small Vessel Disease. Neurology. 2022;99(22):e2454–e2463. doi:10.1212/WNL.0000000000201205

45. UK Biobank. UK Biobank Touch-Screen Fluid Intelligence Test. UK Biobank; 2012. Accessed January 6, 2025. https://biobank.ndph.ox.ac.uk/crystal/crystal/docs/Fluidintelligence.pdf

46. Raulin AC, Doss SV, Trottier ZA, Ikezu TC, Bu G, Liu CC. ApoE in Alzheimer’s disease: pathophysiology and therapeutic strategies. Molecular Neurodegeneration. 2022;17(1):72. doi:10.1186/s13024-022-00574-4

47. Rohn TT. Is apolipoprotein E4 an important risk factor for vascular dementia? Int J Clin Exp Pathol. 2014;7(7):3504–3511.

48. Real R, Martinez-Carrasco A, Reynolds RH, et al. Association between the LRP1B and APOE loci and the development of Parkinson’s disease dementia. Brain. 2023;146(5):1873–1887. doi:10.1093/brain/awac414

49. Tunold JA, Geut H, Rozemuller JMA, et al. APOE and MAPT Are Associated With Dementia in Neuropathologically Confirmed Parkinson’s Disease. Front Neurol. 2021;12:631145. doi:10.3389/fneur.2021.631145

50. Wasserstein RL, Lazar NA. The ASA Statement on p-Values: Context, Process, and Purpose. The American Statistician. 2016;70(2):129–133. doi:10.1080/00031305.2016.1154108

51. Jolma CD, Samson RA, Klewer SE, Donnerstein RL, Goldberg SJ. Acute Cardiac Effects of Nicotine in Healthy Young Adults. Accessed October 1, 2025. https://onlinelibrary.wiley.com/doi/10.1046/j.1540-8175.2002.00443.x

52. Saunders GRB, Wang X, Chen F, et al. Genetic diversity fuels gene discovery for tobacco and alcohol use. Nature. 2022;612(7941):720–724. doi:10.1038/s41586-022-05477-4

53. Sanderson E, Spiller W, Bowden J. Testing and correcting for weak and pleiotropic instruments in two-sample multivariable Mendelian randomization. Stat Med. 2021;40(25):5434–5452. doi:10.1002/sim.9133

54. Elsonbaty SM, Ismail AFM. Nicotine encourages oxidative stress and impairment of rats’ brain mitigated by *Spirulina platensis* lipopolysaccharides and low-dose ionizing radiation. Archives of Biochemistry and Biophysics. 2020;689:108382. doi:10.1016/j.abb.2020.108382

55. Tamagno E, Guglielmotto M, Aragno M, et al. Oxidative stress activates a positive feedback between the γ- and β-secretase cleavages of the β-amyloid precursor protein. J Neurochem. 2008;104(3):683–695. doi:10.1111/j.1471-4159.2007.05072.x

56. Tan JL, Li QX, Ciccotosto GD, et al. Mild oxidative stress induces redistribution of BACE1 in non-apoptotic conditions and promotes the amyloidogenic processing of Alzheimer’s disease amyloid precursor protein. PLoS One. 2013;8(4):e61246. doi:10.1371/journal.pone.0061246

57. Chen X, Guo C, Kong J. Oxidative stress in neurodegenerative diseases. Neural Regen Res. 2012;7(5):376–385. doi:10.3969/j.issn.1673-5374.2012.05.009

58. Heldt NA, Reichenbach N, McGary HM, Persidsky Y. Effects of Electronic Nicotine Delivery Systems and Cigarettes on Systemic Circulation and Blood-Brain Barrier: Implications for Cognitive Decline. The American Journal of Pathology. 2021;191(2):243–255. doi:10.1016/j.ajpath.2020.11.007

59. Heldt NA, Seliga A, Winfield M, et al. Electronic cigarette exposure disrupts blood-brain barrier integrity and promotes neuroinflammation. Brain Behav Immun. 2020;88:363–380. doi:10.1016/j.bbi.2020.03.034

60. Sharma S, Rahman Archie S, Kanchanwala V, et al. Effects of Nicotine Exposure From Tobacco Products and Electronic Cigarettes on the Pathogenesis of Neurological Diseases: Impact on CNS Drug Delivery. Front Drug Deliv. 2022;2. doi:10.3389/fddev.2022.886099

61. Hawkins BT, Abbruscato TJ, Egleton RD, et al. Nicotine increases in vivo blood-brain barrier permeability and alters cerebral microvascular tight junction protein distribution. Brain Res. 2004;1027(1-2):48–58. doi:10.1016/j.brainres.2004.08.043

62. Sweeney MD, Sagare AP, Zlokovic BV. Blood–brain barrier breakdown in Alzheimer’s disease and other neurodegenerative disorders. Nat Rev Neurol. 2018;14(3):133–150. doi:10.1038/nrneurol.2017.188

63. Singh N, Wanjari A, Sinha AH. Effects of Nicotine on the Central Nervous System and Sleep Quality in Relation to Other Stimulants: A Narrative Review. Cureus. 2023;15(11):e49162. doi:10.7759/cureus.49162

64. Jaehne A, Loessl B, Bárkai Z, Riemann D, Hornyak M. Effects of nicotine on sleep during consumption, withdrawal and replacement therapy. Sleep Medicine Reviews. 2009;13(5):363–377. doi:10.1016/j.smrv.2008.12.003

65. Harenbrock J, Holling H, Reid G, Koychev I. A meta-analysis of the relationship between sleep and β-Amyloid biomarkers in Alzheimer’s disease. Biomarkers in Neuropsychiatry. 2023;9:100068. doi:10.1016/j.bionps.2023.100068

66. Whitehead AK, Erwin AP, Yue X. Nicotine and Vascular Dysfunction. Acta Physiol (Oxf). 2021;231(4):e13631. doi:10.1111/apha.13631

67. Venkat P, Chopp M, Chen J. Models and mechanisms of vascular dementia. Exp Neurol. 2015;272:97–108. doi:10.1016/j.expneurol.2015.05.006

68. Mossanen Parsi M, Duval C, Ariëns RAS. Vascular Dementia and Crosstalk Between the Complement and Coagulation Systems. Front Cardiovasc Med. 2021;8:803169. doi:10.3389/fcvm.2021.803169

69. Ben-Shlomo Y, Darweesh S, Llibre-Guerra J, Marras C, Luciano MS, Tanner C. The epidemiology of Parkinson’s disease. Lancet. 2024;403(10423):283–292. doi:10.1016/S0140-6736(23)01419-8

70. van Alten S, Domingue BW, Faul J, Galama T, Marees AT. Reweighting UK Biobank corrects for pervasive selection bias due to volunteering. Int J Epidemiol. 2024;53(3):dyae054. doi:10.1093/ije/dyae054

71. Pagano G, Ferrara N, Brooks DJ, Pavese N. Age at onset and Parkinson disease phenotype. Neurology. 2016;86(15):1400–1407. doi:10.1212/WNL.0000000000002461

72. Valentine G, Sofuoglu M. Cognitive Effects of Nicotine: Recent Progress. Curr Neuropharmacol. 2018;16(4):403–414. doi:10.2174/1570159X15666171103152136

73. Wang H, Sun X. Desensitized nicotinic receptors in brain. Brain Research Reviews. 2005;48(3):420–437. doi:10.1016/j.brainresrev.2004.09.003

74. Kim SH, Yun CH, Lee SY, Choi KH, Kim MB, Park HK. Age-dependent association between cigarette smoking on white matter hyperintensities. Neurol Sci. 2012;33(1):45–51. doi:10.1007/s10072-011-0617-1

75. Lousdal ML. An introduction to instrumental variable assumptions, validation and estimation. Emerg Themes Epidemiol. 2018;15:1. doi:10.1186/s12982-018-0069-7

76. Reed ZE, Wootton RE, Khouja JN, et al. Exploring pleiotropy in Mendelian randomisation analyses: What are genetic variants associated with ‘cigarette smoking initiation’ really capturing? Genet Epidemiol. 2025;49(1):e22583. doi:10.1002/gepi.22583

77. Mayeda ER, Glymour MM, Quesenberry CP, Whitmer RA. Inequalities in dementia incidence between six racial and ethnic groups over 14 years. Alzheimers Dement. 2016;12(3):216–224. doi:10.1016/j.jalz.2015.12.007

78. Derby KS, Cuthrell K, Caberto C, et al. Nicotine Metabolism in Three Ethnic/Racial Groups with Different Risks of Lung Cancer. Cancer Epidemiol Biomarkers Prev. 2008;17(12):3526–3535. doi:10.1158/1055-9965.EPI-08-0424

79. Benowitz NL, Perez-Stable EJ, Fong I, Modin G, Herrera B, Jacob P. Ethnic differences in N-glucuronidation of nicotine and cotinine. J Pharmacol Exp Ther. 1999;291(3):1196–1203.

80. Djordjevic MV, Stellman SD, Zang E. Doses of Nicotine and Lung Carcinogens Delivered to Cigarette Smokers. J Natl Cancer Inst. 2000;92(2):106–111. doi:10.1093/jnci/92.2.106

81. Frederiksen LW, Miller PM, Peterson GL. Topographical components of smoking behavior. Addictive Behaviors. 1977;2(1):55–61. doi:10.1016/0306-4603(77)90009-0

82. Xing Y, Yang J, Zhou A, et al. White Matter Fractional Anisotropy Is a Superior Predictor for Cognitive Impairment Than Brain Volumes in Older Adults With Confluent White Matter Hyperintensities. Front Psychiatry. 2021;12:633811. doi:10.3389/fpsyt.2021.633811

83. Raghavan S, Przybelski SA, Reid RI, et al. Reduced fractional anisotropy of the genu of the corpus callosum as a cerebrovascular disease marker and predictor of longitudinal cognition in MCI. Neurobiology of Aging. 2020;96:176–183. doi:10.1016/j.neurobiolaging.2020.09.005

84. Raghavan S, Reid RI, Przybelski SA, et al. Diffusion models reveal white matter microstructural changes with ageing, pathology and cognition. Brain Commun. 2021;3(2):fcab106. doi:10.1093/braincomms/fcab106

85. Wang M, Liu D, Yang S, Li Y, Lian X. Smoking, alcohol consumption, and age at onset of Huntington’s disease: a Mendelian randomization study. Parkinsonism & Related Disorders. 2022;97:34–38. doi:10.1016/j.parkreldis.2022.02.013

86. Goodheart AE, Gomperts SN. The association between cigarette smoking and dementia with Lewy bodies. Parkinsonism & Related Disorders. 2024;128:107133. doi:10.1016/j.parkreldis.2024.107133

87. Lawton M, Ben-Shlomo Y, Gkatzionis A, Hu MT, Grosset D, Tilling K. Two sample Mendelian Randomisation using an outcome from a multilevel model of disease progression. Eur J Epidemiol. 2024;39(5):521–533. doi:10.1007/s10654-023-01093-2

88. Lovegrove CE, Howles SA, Furniss D, Holmes MV. Causal inference in health and disease: a review of the principles and applications of Mendelian randomization. J Bone Miner Res. 2024;39(11):1539–1552. doi:10.1093/jbmr/zjae136

89. UK Biobank. UK Biobank Research Ethics Approval. UK Biobank; 2021. Accessed March 6, 2025. https://www.ukbiobank.ac.uk/learn-more-about-uk-biobank/about-us/ethics

